# Magnetic bead-based ELISA allow inexpensive, rapid and quantitative detection of human antibodies against SARS-CoV-2

**DOI:** 10.1101/2020.07.26.20162255

**Authors:** Luciano Fernandes Huergo, Marcelo Santos Conzentino, Edileusa Cristina Marques Gerhardt, Adrian Richard Schenberger Santos, Fabio Oliveira Pedrosa, Emanuel Maltempi Souza, Meri Bordignon Nogueira, Karl Forchhammer, Fabiane Gomes Moraes Rego, Sônia Mara Raboni, Rodrigo Arantes Reis

**Affiliations:** Setor Litoral, UFPR Matinhos, PR, Brazil; Departamento de Bioquímica e Biologia Molecular, UFPR Curitiba, PR, Brazil; Complexo Hospital das Clínicas, UFPR Curitiba, PR, Brazil; Interfakultäres Institut für Mikrobiologie und Infektionsmedizin der Eberhard-Karls Universität Tübingen, Germany; Post-Graduation Program in Pharmaceutical Sciences, Federal University of Paraná, Curitiba, PR, Brazil, UFPR Curitiba, PR, Brazil; Departamento de Biologia Celular, UFPR Curitiba, PR, Brazil

**Keywords:** SARS-CoV-2, COVID-19, immunological test, ELISA, magnetic beads

## Abstract

Here we describe a novel immunogenic method to detect COVID-19. The method is a chromogenic magnetic bead-based ELISA which allows inexpensive and quantitative detection of human IgG or IgM antibodies against SARS-CoV-2 in serum or whole blood samples in just 12 minutes. As a proof of concept, we compared the performance of our new method to classical ELISA. Person correlation between optical densities obtained using the two methods was 0.98, the color intensity observed in the novel method correlated with antibody titers determined by classical ELISA. The novel magnetic bead-based ELISA correctly classified all 6 positive COVID-19 samples tested and showed 100% specificity as judged by the analysis of a cohort of 26 negative samples. The magnetic bead-based ELISA performed better than classic ELISA to discriminate COVID-19 positive serum with low antibody titer. The chromogenic magnetic bead-based ELISA method described here can be applied to both point of care and high throughput analysis. The method is readily adaptable to be used with other protein and peptide-based antigens.

## Introduction

In December 2019, health authorities in Wuhan, China, reported cases of patients with pneumonia of unknown epidemiological causes, linked to a seafood/animal market. The pathogen in these cases has been identified, through viral isolation, electron microscopy and RNA sequencing, as a new beta-coronavirus called SARS-CoV-2 (Zhu *et al*. 2020). Shortly after its emergence in China, SARS-CoV-2 spread rapidly across the world. According to the John Hopkins University website (https://coronavirus.jhu.edu/map.html), on 18/07/2020, more than 14 million cases of COVID-19 have been registered worldwide and the disease was responsible for 600,000 deaths. So far, there is no vaccine or drug to treat the virus and/or the disease. The only effective measure is the isolation of infected subjects and social distance, which, in turn, has profound socioeconomic impacts specially in less developed countries such as Brazil.

The most effective strategy for combating COVID-19 is to conduct massive diagnostic tests and quickly isolate infected subjects. The widely used qRT-PCR has been considered the gold standard molecular method for detecting SARS-CoV-2 RNA from swab samples. The shortcoming of qRT-PCR is that it is limited to track virus during the acute phase of the disease. Furthermore qRT-PCR is expensive and requires sophisticated instrumentation, well trained personnel, intensive labor and is not adaptable to point of care analysis (Petherick 2020). This is particularly critical to countries such as Brazil where samples may need to travel long distances between patients and analytical laboratories adding additional transportation costs. Furthermore, samples may deteriorate during transport if not handle properly.

In contrast to qRT-PCR, COVID-19 immunological tests detect antibodies that react against SARS-COV-2 antigens in human serum samples (Petherick 2020). One of the main advantages of immunological tests is that they can be used to screen for COVID-19 cases after the acute phase of the disease, since the antibodies titer remains detectable in symptomatic and asymptomatic cases even after 8 weeks of onset symptoms (Long *et al*. 2020b, 2020a). Hence, COVID-19 immunological tests are important tools for epidemiological surveillance strategies and for analyzing the efficiency of future COVID-19 vaccines.

Usually, the SARS-COV-2 IgG and IgM antibody titers reaches a plateau 11-16 days after symptoms onset (Long *et al*. 2020a), however, some patients produce detectable antibody titers already 2-4 days after symptoms onset (Long *et al*. 2020b, 2020a). Thus, COVID-19 immunological tests may also be useful as an additional toolkit to identify patients in the COVID-19 acute phase, even those who tested as false negative in qRT-PCR (Long *et al*. 2020a).

The ELISA assay (*enzyme-linked immunosorbent assay*), described by Engvall and Perlmann in the early seventies (Engvall and Perlmann 1972), is still considered the gold standard immunological method as it can provide a relatively low-cost, easy to implement and high-throughput method to detect antibodies quantitatively. It still requires relatively expensive instrumentation, trained personnel and about a day to complete the analysis. Furthermore, ELISA is not suitable to point of care analysis. On the other hand, lateral flow immunochromatography, which has been widely used to track COVID-19 cases, is relatively inexpensive and can be applied at point of care providing the results typically in 20 min (Carter *et al*. 2020). The shortcomings of immunochromatography tests are that it provides only qualitative data, it is not adaptable to high-throughput and the accuracy of some of the tests available on the market has been questioned (Adams *et al*. 2020).

Here we describe a chromogenic magnetic bead-based ELISA, which allows inexpensive, quantitative, and scalable detection of human antibodies against SARS-CoV-2 in 12 minutes. The method can be easily adjusted to point of care or to the high throughput format requiring minimal personnel training and using only 2 µl of blood as biological material. The results can be interpreted by visual inspection without instrumentation. The estimated cost per test is 1 US$ and the entire process from antigen production to the results data can be implemented with minimal lab instrumentation.

## Material and Methods

### Expression and purification of recombinant SARS-CoV-2 Nucleocapsid protein (N-protein)

A codon optimized synthetic gene expressing the full-length SARS-CoV-2 N-protein (QHD43423.2) was obtained and cloned into pET28a by General Biosystems. This plasmid was named pLHSarsCoV2-N and transformed into *E. coli* BL21 (λDE3) enabling the expression of N-terminal His-tagged N-protein after induction with IPTG. The cells were growth in 400 ml LB medium at 120 rpm at 37°C to OD_600nm_ of 0.3. Then, IPTG was added to a final concentration 0.3mM and 30 min later, the incubator temperature was lowered to 16°C. The culture was kept at 120 rpm at 16°C over/night. Cells were collected by centrifugation at 3,000 xg for 5 min. The cell pellet was resuspended in 10 ml of buffer 1 (Tris-HCl pH 8 50mM, KCl 100 mM, imidazole 20 mM). Cells were disrupted by sonication on an ice bath. The soluble fraction was recovered after centrifugation at 20,000 xg for 10 minutes and loaded onto a 1 ml Histrap chelating Ni^2+^ column (GE Healthcare), which had been previously equilibrated with buffer 1. The column was washed with 10ml of buffer 1 and bound proteins were eluted with buffer 1 containing increasing concentrations of imidazole. The His-tagged N-protein eluted at 300 mM. The fractions were pooled and desalted using a 5 ml desalting column (GE Healthcare) equilibrated with 50 mM Tris-HCl pH 8, 100 mM KCl and 10% (v/v) glycerol as buffer. The final protein preparation yield was 3 mg and was homogeneous as judged by SDS-PAGE analysis (Fig. S1). The purified protein was stored in aliquots at 4°C for one week or at −20°C for up to two months. The protein identity was confirmed by peptide mass fingerprint MALDI-TOF analysis (Fig. S1).

### Human sera

Samples were collected from patients at the Complexo Hospital de Clínicas (CHC/UFPR), a tertiary academic hospital. Covid-19 cases were confirmed by the detection of SARS-CoV-2 RNA by real-time RT-PCR from nasopharyngeal sample swabs by virology laboratory. The time point of sampling of serum ranging from 3 to 30 from the onset symptoms. Negative samples were obtained from blood donors bank collected from healthy individuals in 2018. The Institutional Ethics Review Board of CHC/UFPR (n# 30578620.7.0000.0008) approved this study. Human sera were inactivated at 56°C for 30 minutes before the analysis.

### ELISA assays

The purified SARS-CoV-2 N-protein was diluted to 2 ng/µl in 50 mM Tris-HCl pH 8, 100 mM KCl, 10% (v/v) glycerol and 0.1 ml aliquots were transferred wells of ELISA 96 well plates (Olen) overnight at 4°C. The next day, the unbound excess protein was removed, and wells were washed twice with 0.2 ml of 1x TBST buffer. Wells were blocked with 0.2 ml 3% (w/v) skimmed milk in 1x TBST overnight at 4°C. The serum was diluted in 1x TBST, 1% (w/v) skimmed milk (the dilution factors are indicated in each experiment) and 0.1 ml were loaded onto the ELISA wells following by incubation at room temperature for 1 hour. The wells were washed three times with 0.2 ml of 1x TBST followed by incubation with 0.1 ml of anti-human IgG-HPR from goat (Thermo Scientific) diluted 1:3000 in 1x TBST. Wells were washed three times with 0.2 ml of 1x TBST. The ready to go HPR substrate TMB (Thermo Scientific) was added to the wells (0.1 ml). The reaction was stopped after 10 min incubation at room temperature by addition of 0.1 ml 1M HCl. The plates were put on the top of a white light transilluminator device and photographed. The optical density was measured at 450 nm using a TECAN Nano plate reader (TECAN).

### Magnetic beads-based ELISA

The magnetic bead-based ELISA was performed using MagneHis Ni^2+^ magnetic particles (Promega). Ten micro liters of beads were washed in 0.5 ml of 1x TBST and resuspended in 0.5 ml of 1x TBST. Ten µg of purified His-tagged N-protein was incubated with the washed magnetic beads for 5 min at room temperature with gentle mixing. The beads were washed once with 0.5 ml of 1x TBST containing 1% (w/v) skimmed milk and resuspended in 1.2 ml of the same wash buffer, 0.1 ml were distributed on each well of a 96 well plate (Fig. 1). Two micro liters of human serum were diluted in 0.2 ml of TBST 1x skimmed milk 1% (w/v) directly on the wells of the 96 well plate. The magnetic beads coated with N protein were mixed with the diluted serum for 2 min. The beads were captured using a simple custom-made magnetic extractor device (see below) and loaded into sequential 2x wash steps for 30 seconds in 1x TBST. The beads were incubated for 2 min with 0.15 ml goat anti-human IgG-HPR (Thermo Scientific) diluted 1:3000 in 1x TBST, following by 2x wash steps for 30 seconds TBST 1x. The beads were transferred to wells with 0.15 ml of the HPR substrate TMB (Thermo Scientific) and 5 min. The total time of the procedure was typically 12 minutes. All steps were performed at room temperature. We routinely performed the test in 12 samples format (Fig. 1). When the reactions were complete the beads were removed, the plates were put on the top of a white light transilluminator device and photographed. The optical density was measured at 650 nm using a TECAN Nano plate reader (TECAN).

**Fig 1.**
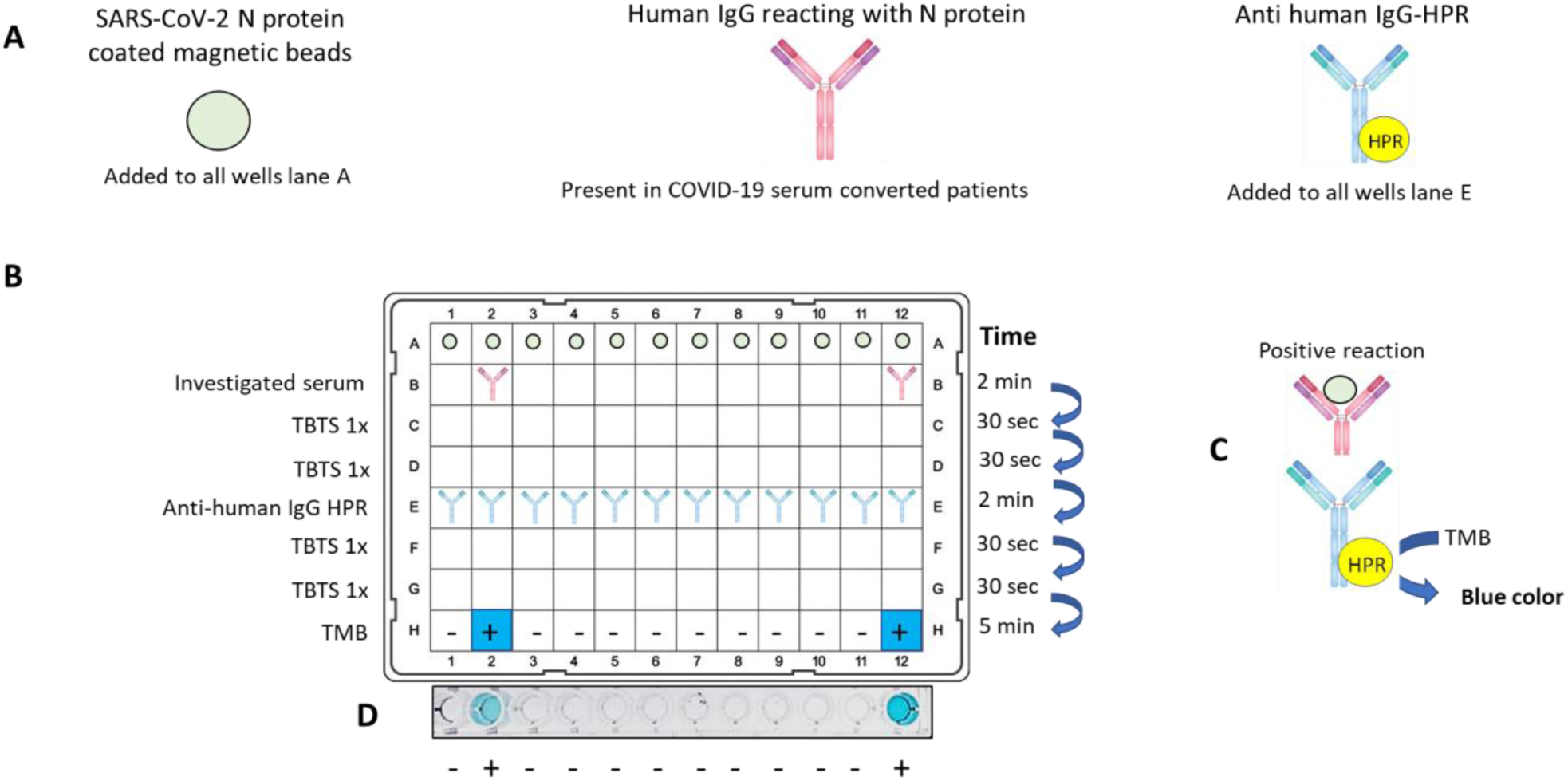
Magnetic bead-based chromogenic ELISA. A) Reaction components. B) Diagram of the reaction and distribution of the components on a 96 well plate. C) Positive samples will develop a blue color in line H due to the formation of a ternary complex between antigen, primary and HPR secondary antibody. D) Photography register of a typical assay. H2 and H12, serum from two PCR-confirmed patients (mild and severe, respectively). Other lanes serum from true negative subjects collected in 2018.

### Magnetic bead extractor device

A homemade magnetic extractor device was prepared fixing 12 nails (11mm) on a piece of foam (Fig. S2A). In the head of the nails a set of neodymium magnets were added by simple magnetic attraction. We used 2x (3mm diameter × 2 mm) and 1x (2 mm × 2 mm) neodymium magnets (Fig. S2A). A better-quality system can be prepared using a 3D printer (Fig. S2B) (the file for printer is available upon request). The extraction of the beads is performed by inserting the magnets on the PCR-strip so the PCR-strip will hold firmly the magnetic beads during the transfer from well to well (Fig. S2). Once the beads are in place in the next solution, magnets are removed, and the PCR strip is gently moved to allow proper homogenization of the magnetic beads in the solution. The time of each step is recorded once the beads turn homogeneous.

## Results

The aim of this work was to develop an inexpensive COVID-19 immunological rapid test, adaptable to point of care analysis, which would provide not only qualitative but also quantitative data using low volume of biological sample. The basic idea was to develop an indirect chromogenic ELISA assay, which proceeds not as usual with the antigen immobilized on the surface of a polystyrene plate but on the surface of a magnetic bead nanoparticle. We predicted that the use of nanoparticles would result in increased surface area of exposed antigens allowing a faster kinetics of protein association / dissociation during the process thereby allowing the reduction of incubation times and unspecific background.

As a proof of concept, we expressed and purified an N-terminal His-tagged version of SARS-CoV-2 Nucleocapsid N protein and immobilized it onto commercially available nickel magnetic particles. These particles were then challenged with human serum from COVID-19 positive and negative patients for two minutes with the procedure described in Fig. 1. The beads were extracted from the solution using a homemade inexpensive magnetic beads extractor device and transferred to two sequential 30 seconds washes in TBST buffer. Beads were then immersed in secondary conjugated anti-human IgG HPR for 2 minutes, which was followed by two sequential 30 seconds washes in TBST buffer. The beads were finally immersed in TMB chromogenic substrate and incubated for 5 minutes to develop the blue color. Beads were removed and the result can be interpreted by visual inspection. The samples containing COVID-19 negative sera were completely blank while those from two COVID-19 patients (one mild and one severe convalescent subject) developed a blue color, which indicated the presence of IgG antibodies recognizing the SARS-CoV-2 N protein (Fig. 1). When all reagents are already in place, the procedure takes about 12 minutes and uses only two micro liters of serum as start material.

To confirm the efficiency of our novel immunological method, we compared the results with classical ELISA, which is considered the gold standard immunological method. Classical ELISA was in house built using the same antigen preparations (the SARS-CoV-2 N protein) mounted on polystyrene 96 well plates. Serum from a COVID-19 positive patient (PCR-confirmed) and a negative control (collected in 2018) serum were subjected to serial dilutions and allowed to react with SARS-CoV-2 N protein on both classic ELISA and magnetic particles-based ELISA. On regular ELISA, the COVID-19 positive serum showed strong reaction with the N protein in dilution for up to 1:25,600 (Fig. 2A). A clearly distinguished profile was observed in the negative control serum, which showed background cross reaction only in dilutions up to 1:800 (Fig. 2A). When the same samples were analyzed in our magnetic bead-based ELISA, the COVID-19 positive serum showed strong reaction with the N protein antigen in dilutions for up to 1:3,200. On the other hand, the negative control serum had no detectable background cross reaction even at low dilutions (Fig. 2B). These data show that magnetic bead-based ELISA provided quantitative data and can therefore be used to determine serum titers without background cross reactions characteristic for classic ELISA. The linear range of quantification was between 400 to 25,600 serum dilution fold with R^2^ = 0.995 (Fig. 2C).

**Fig 2.**
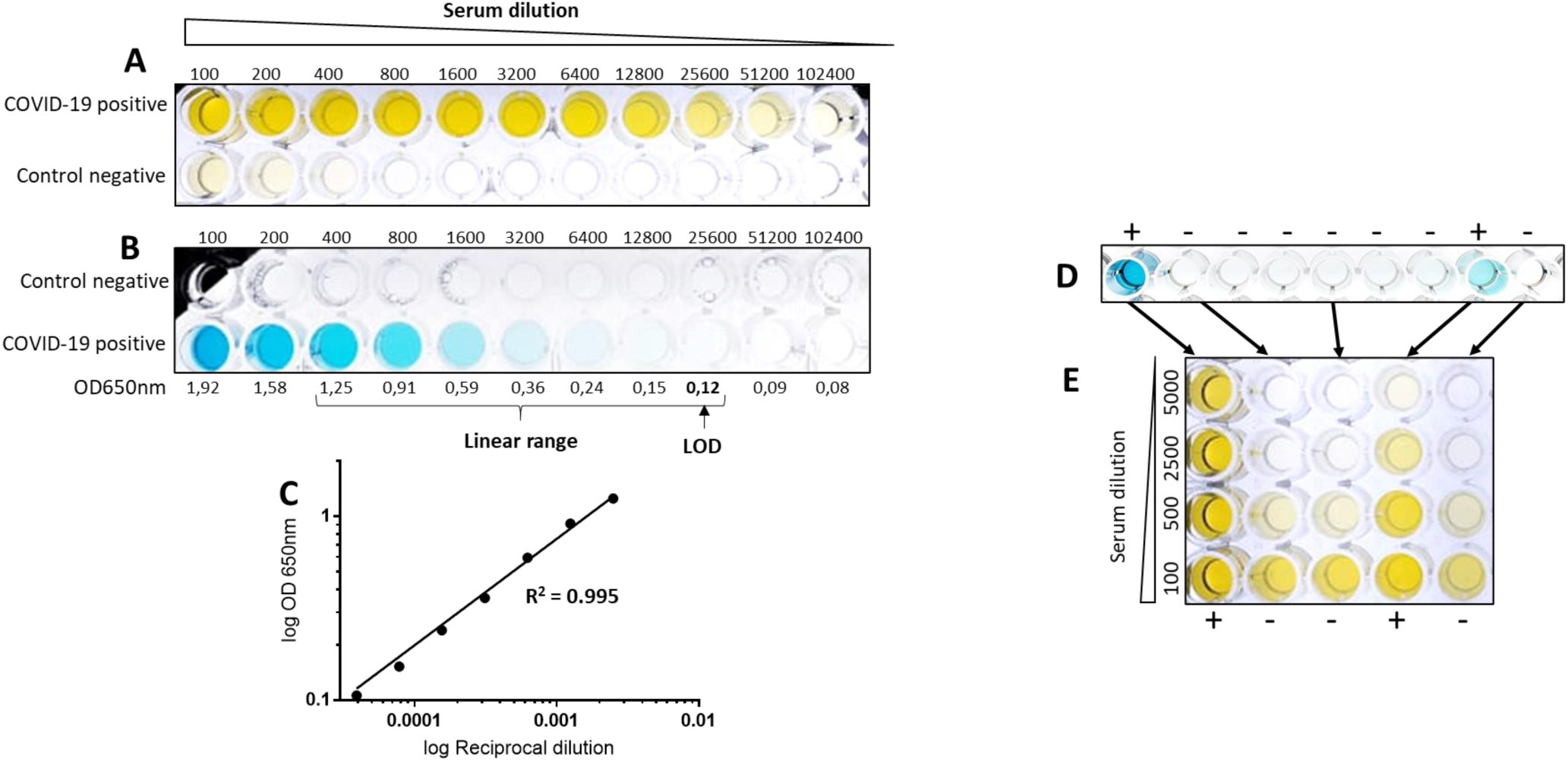
Serum dilution titer in regular ELISA and Magnetic-bead-based ELISA. The serum from a COVID-19 coalescent subject and a control negative subject were serial diluted and allowed to react with SARS-CoV-2 N protein coated regular ELISA (A) or magnetic beads-based ELISA (B). The limits of detection (LOD) was determined as 2x S/N ratio. C) Log scale regression of OD650nm vs reciprocal serum dilution, data from rapid test in figure B. The linear range for quantification was from 400 to 25600 dilution with R^2^ = 0.995. The serum from a COVID-19 positive subjects (+) and a control negative subjects (-) were analyzed using magnetic beads-based ELISA analysis (D) and by classic ELISA at indicated dilutions (E). The titer of the serum observed in classical ELISA clearly correlate with the color developed on the magnetic bead-based ELISA.

The absence of background in negative samples using magnetic bead-based ELISA is likely to be a consequence of the shorter incubation time with the primary antibody, which allows only the effective capture of high affinity antibodies from the serum. Furthermore, the increased surface area in the magnetic beads system in comparison to the plastic surface improves the efficiency of the washing steps removing low affinity antibodies interactions. In fact, only two washes were required to provide zero cross reactive background in the negative control samples. Conversely, in our regular ELISA, three washing steps were applied, and we still detected cross reactivity at low serum dilutions (Fig. 2A).

The correlation between classic ELISA and Magnetic bead-based ELISA was further evaluated by comparing the color developed in the magnetic system versus serum titration on classical ELISA. Two PCR-confirmed COVID-19 cases (severe and mild) were analyzed along with a set of true negative samples (collected in 2018). Negative samples were blank in magnetic bead system (Fig. 2D) whereas mild and severe COVID-19 samples showed development of blue color with color intensity clearly correlated with the serum titer observed using classic ELISA (Fig. 2D and E). The COVID-19 positive and negative samples could be easily discriminated by visual inspection (Fig. 2D).

The robustness of the assay was further investigated with a set of six PCR-confirmed cases along with 6 real negative samples which were analyzed using magnetic bead system and classic ELISA. Again, the color developed in both methods clearly correlate, the Person correlation of the optical density was 0.98 (Fig. 3A and B). One of the PCR-positive samples (number 26) had little antibody titer as indicated by the faint blue color developed (Fig. 3A). This positive case was difficult to differentiate from the negative samples by simple visual inspection. When the optical density was measured, sample 26 could be classified as a true positive using magnetic bead ELISA but not when using classical ELISA (Fig. 3B and C). Hence, the magnetic bead-based ELISA perform better than classical ELISA especially for low antibody titer samples. The novel magnetic bead-based ELISA correctly classified all 6 positive COVID-19 samples tested and showed 100% specificity as judged by the analysis of a cohort of 26 negative samples.

**Fig 3.**
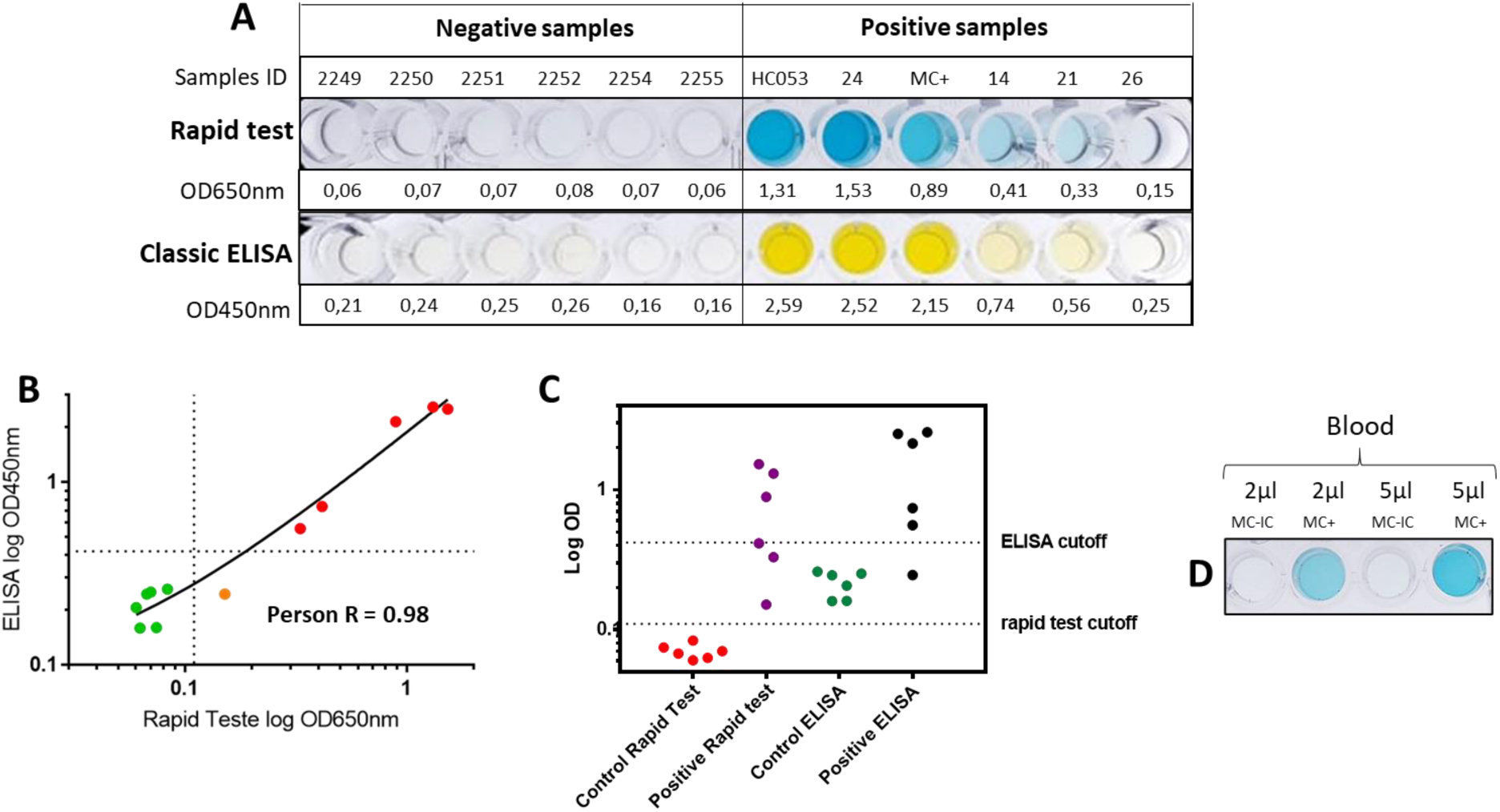
Comparison between classic and Magnetic-bead-based ELISA. A) The serum from Negative samples (collected in 2018) and COVID-19 PCR positive samples were compared using the rapid test (1:100 dilution) and Classical ELISA (1:1,000 dilution). B) Person correlation between OD registered in classical vs magnetic bead-based ELISA. Negative subjects are in green dots. Positive in red. In orange is serum from a PCR positive subject which was considered positive by magnetic bead ELISA but as negative by classical ELISA. C) Log OD Distribution recorded for the different groups of samples. The cutoff line for ELISA and rapid test was determined as mean value of negative samples + 5x SD. D) Whole blood can be directly applied detect COVID-19 positive cases using magnetic bead ELISA. The indicated amount of fresh blood from a negative subject (MC-IC, subject who did not have contact with COVID-19 positive subjects nor have any of the COVID-19 related symptoms since December 2029) and positive subject (MC+, who had mild synthons and tested PCR positive 38 days before blood sampling) was directly used for the analysis.

We evaluated if whole blood could be used instead of serum in our Magnetic bead-based ELISA. Fresh blood (2 or 5 µl) were collected from a healthy subject and from a COVID-19 convalescent subject and immediately diluted in 0.2 ml of TBST 1x skimmed milk 1% (w/v) on the wells of the 96 well plate. The assay was performed exactly was described in Fig. 1, except that whole blood instead of serum was applied on the wells of line B. The data in Fig. 3D indicate that only 2 µl of whole blood can be used as star biological material. Again, as no cross-reaction background was observed in the negative control and the positive sample developed an intense blue color, results could be visually inspected. These data further support that the method is well suited for point of care analysis.

## Discussion

Here we describe a novel Magnetic bead-based chromogenic ELISA which allowed rapid and quantitative detection of antibodies against SARS-CoV-2 (Fig. 1). As a proof of concept, the performance of this novel method was compared to data obtained when the same samples were analyzed using classical ELISA. We noted that while COVID-19 negative serum showed some cross reactivity with N protein antigen using classical ELISA under low serum dilutions, such cross reaction was not detectable in the Magnetic bead-based ELISA (Fig. 2). Hence, the Magnetic bead-based ELISA can be used to separate COVID-19 positive and negative serum by visual inspection of the results without the need of instrumentation (indeed we just had access to and microplate reader in the end of this project). The method showed similar results when whole blood was used as starting biological material (Fig. 3D). Therefore, our method can be readily adaptable to point of care analysis. The color developed in the Magnetic beads-based ELISA clearly correlated with the antibody titers obtained in classical ELISA (Fig. 2 and 3), thus, our novel method can provide quantitative antibody data in 12 minutes.

In this optimized system, we used 1 µg of N protein antigen and 1 µl of magnetic beads per test. We estimated that the total price of consumables per sample tested to be approximately R$ 5 (about $1 US). The price can be further reduced with the use of minimal autoinduction medium for the expression of the SARS-CoV-2 N protein antigen, the purification of the protein antigen directly on the Ni^2+^ magnetic beads and also with the homemade preparation of TMB.

We optimized the reaction system to use only 2 µl of human serum and incubate the N-protein coated magnetic particles with the serum for 2 min. We noted that the incubation time, the amount of serum and the time for color development can be increased, thus enhancing the color signal of the reaction. However, this may increase cross reactivity background in the negative control serum. Hence, adjustment of reagents concentrations and incubation time are critical to obtain data which can be interpreted visually. The presence of EDTA, which is commonly used as an anticoagulant in serum, did not affect the performance of the method in concentrations up to 50 µM (Fig S3A). The method can be easily adjusted for IgM analysis just by changing the secondary antibody (Fig. S3B).

The Magnetic bead-based ELISA described here can be readily adapted to use any other 6x His tagged protein as antigen for COVID-19 or to detect other antibodies for studying other diseases. Furthermore, other commercial magnetic resins and protein affinity tags may be used (i.e streptavidin magnetic beads with Strep-tag or biotinylated antigens; anti-FLAG magnetic beads with FLAG tagged antigens). Currently, we are using the format described in Fig. 1A with 12 samples being run simultaneously and delivering the results in 12 minutes. However, the whole process can be scaled up to high throughput using a full 96 well plate for each step and commercially available manual or automatic magnetic extractor devices.

In a recent review of different serological techniques for COVID-19 diagnostics (Carter *et al*. 2020), all methods described required much longer time to deliver results than the novel method we describe here. There is only one report of a synthetic peptide-coated magnetic-bead chemiluminescence ELISA, which provides 71% sensitive for COVID-19 IgG (Cai *et al*. 2020). In the work by Cai *et al*., biotinylated synthetic peptides comprising different parts of SARS-CoV-2 proteins were bound to streptavidin-coated magnetic beads. The assays used 0.1 ml of serum as starting material. The total time of the overall process has not been described, however, only the first incubation step already took 10 minutes and a total of ten washing steps was required before the results were delivered by chemiluminescence. This method is available on the market (Bioscience), which claims to deliver results in 30 min.

We consider that our method described here has several advantages in comparison to those described by Cai *et al*.; 1) Much less serum is required as start material, so a simple lancet can be used to collect blood instead of venous puncture; 2) Whole blood can be used, only 2 µl showed good performance as zero background in negative subjects (Fig. 3D). 3) Reactions can be interpreted by visual inspection, which facilitates point of care analysis; 4) The total reaction time is at least 2.5 × shorter; 5) The antigen can be produced at large scale with very low cost and without requirement of sophisticated lab instrumentation. It is worth mentioning that in the work by Cai *et al*., several important methodological information was missing (i.e details of peptides and buffers) making it impossible to reproduce the process.

Several different indirect ELISA kits are now available commercially for COVID-19 diagnostic. Recent literature data showed that homemade indirect ELISA kits perform well in terms of sensitivity and specificity to detect COVID-19 cases when using either Spike or Nucleocapsid protein as antigen (Amanat *et al*. 2020; OKBA *et al*. 2020). Considering that we could show good agreement between our magnetic bead-based ELISA and classic ELISA across all samples used in this study (Fig. 2 and 3), we speculate that our method will perform similar or even better than classical ELISA in terms of specificity and sensitivity. Despite the small cohort analyzed in this study (6 positive and 26 negative samples), our method was able to clearly separate all COVID-19 positive and negative cases in minutes. Analysis of larger cohort of samples are underway to provide better statistics of specificity and sensitivity. It worth mentioning that the use of a microplate reader may improve the performance of the assays specially for positive cases carrying low antibody titer (Fig. 3)

We believe that the inexpensive, rapid, and quantitative method to detect human antibodies against SARS-CoV-2 described in this study may help to track COVID-19 cases specially in developing countries such as Brazil. The assay requires minimum instrumentation in all phases of production, and it is ready to be evaluated as a COVID-19 diagnostic toll with larger sample cohort and to scale-up production. The technology is easily adaptable for diagnosis of other diseases.

## Data Availability

Data is available upon request

## Acknowledgments

This work was supported by CNPq, UFPR, Fundação Araucária, CAPES and by the Alexander von Humboldt foundation. We would like to thank the technical assistance of Felipe F. Souza and Mariana Nazário. We are grateful to Tatielle Pricila Cintra dos Santos (UFPR-litoral) for designing and printing of the magnet holder.

